# Inflammatory profile of induced sputum composition in systemic sclerosis: a comparison with healthy volunteers

**DOI:** 10.1101/2020.11.23.20236679

**Authors:** P. Jacquerie, M. Henket, B. André, C. Moermans, D. de Seny, F. Gester, R. Louis, M. Malaise, J. Guiot

## Abstract

**Background:** Systemic sclerosis (SSc) is a potentially serious and disabling connective tissue disease specially in case of interstitial lung disease (SSc-ILD). The aim of our study was to evaluate the potential utility of dosing in the induced sputum (IS) and to compare their levels in SSc-ILD and SSc-nonILD patients, as well as in healthy volunteers (HV). IS and sera values were also compared.

**Methods:** In a prospective cross-sectional analysis, we studied the IS and serum provided from 25 SSc patients, 15 SSc-nonILD and 10 SSc-ILD, compared to 25 HV. We analyzed sputum cell composition and quantified in the supernatant and corresponding serum by commercially available immunoassays: IGFBP-1, IGFBP-2, IGFBP-3, TGF-β, IL-8, TNF-α, YKL-40, MMP-7 and MMP-9. Lung function was studied by the determination of FEV-1 (%), FVC (%), DLCO (%) and KCO (%).

**Results:** The IS of SSc patients had a lower weight than HV (p<0.01) without any significant difference with regard to the cellularity. IGFBP-1 (p<0.0001), TGF-β (p<0.05), IL-8 (p<0.05), YKL-40 (p<0.0001) and MMP-7 (p<0.01) levels were increased in the IS of SSc patients compared to HV. Only IL-8 serum levels (p<0.001) were increased in SSc patients compared to HV. Neither in IS nor in serum were observed differences between SSc-ILD and SSc-nonILD patients. Correlations were observed between IS IL-8 levels and FEV-1 (%)(r=-0.53, p<0.01), FVC (%) (r=-0.51, p<0.01) and annualized □KCO (%) (r=0.57, p<0.05), between IS TGF-□ levels and annualized □FEV-1 (%) (r=-0.57, p<0.05), between IS IGFBP-2 levels and annualized □KCO (%) (r=0.56, p<0.05).

**Conclusion:** Our study showed that SSc patients exhibit raised IS levels of IGFBP-1, TGF-β, IL-8, YKL-40 and MMP-7, molecules known to be involved in lung remodeling and fibrotic process, without any significant difference between SSc-ILD and SSc-nonILD patients. IL-8, TGF-□ and IGFBP-2 are correlated with lung function in SSc patients which emphasize clinical relevance. IS analysis represents a new approach to understand lung inflammatory process in SSc patients. A longitudinal study is needed to evaluate their pathophysiological relevance.

## Introduction

Systemic sclerosis (SSc), also called scleroderma, is a chronic immune-mediated rheumatic disease characterized by fibrosis of the skin and visceral organs (heart, kidneys, lungs and gastrointestinal tract), narrowing of vascular lumen by intimal fibrosis leading to distal ischemia (almost constant Raynaud’s phenomenon), and production of autoantibodies (Ab) such as anti-centromere, anti-topoisomerase 1 (anti-Scl 70) and more rarely anti-RNA polymerase III Abs [1].

Two major subsets of SSc are commonly recognized and are distinguished by the extent of the skin sclerosis: diffuse cutaneous involvement extends beyond the elbows and knees, affecting the proximal limbs and/or the trunk (dcSSc) while in the limited cutaneous form (lcSSc), less than 5% of SSc patients, the injury does not rise above the elbows and knees. In forms without skin involvement, a distinction is also made between (early) limited SSc (most often Raynaud phenomena and specific Abs without visceral damage) and sine scleroderma, a very rare condition where the patient has the internal organ manifestations without the skin findings. The prognosis depends mainly on the presence of visceral involvement and mostly of interstitial lung disease (ILD), pulmonary arterial hypertension (PAH) or renal crisis [2-7]. From an immunological point of view, anti-centromere Abs are related to limited SSc and are more often associated with PAH, anti-topoisomerase I are more related to diffuse SSc and frequently associated with ILD, whereas anti-RNA polymerase III Abs are more often observed in diffuse SSc complicated by renal crisis [8].

ILD is a common complication of systemic sclerosis that is often progressive and has a poor prognosis [9-11]. In contrast with the limited forms, patients with diffuse cutaneous SSc are at increased risk of developing ILD early in the course of their disease [12-13]. High resolution thoracic computed tomography (HRCT) is the gold standard for detecting ILDs [14] in patients with connective tissue diseases. Up to 90% of patients will have interstitial abnormalities on HRCT [15]. Parenchymal lung involvement often appears early after the diagnosis of SSc, with 25% of patients developing clinically significant lung disease within 3 years [16] mainly with a radiological pattern of nonspecific interstitial pneumonia (NSIP)[17]. Pulmonary function test (PFT) can be used to assess the severity of the restrictive syndrome (decrease in total lung capacity <80% predicted)[18]. The alveolo-capillary dysfunction assessed by diffusing capacity of CO (DLCO) appears to be also impaired in fibrotic lung diseases [19]. Due to its invasiveness and its limited contribution, bronchoalveolar lavage fluid (BALF) evaluation is not routinely performed to systematically assess SSc-ILD [20] but used specifically in case of suspected active infection, especially opportunistic ones associated with immunosuppressive therapies. Therefore, induced sputum (IS), which is less invasive and of clinical interest for airway diseases such as idiopathic pulmonary fibrosis (IPF)[21-24], can be proposed as a good non-invasive tool to study lung inflammation in patients with SSc. Previous researches using IS and BALF in SSc have mainly focused on the cellularity [25-27]. If many biomarkers have already been studied both in serum and BALF from ILD patients and in particular in IPF [23,28], very few works have been done in SSc as recently reviewed [29]. We have found only one study suggesting that a lower sputum level of caveolin-1, involved in the pathway of the TGF-β, could be a relevant biomarker for SSc-ILD detection [30].

The aim of our study was to evaluate the potential of measuring parameters of the fibrosing process in sputum from patients with SSc in comparison with healthy volunteers (HV), to compare between SSc-ILD and SSc-nonILD patients, and lastly to compare between IS and serum in all groups. IS levels of biological parameters were also compared to pulmonary function tests in SSc patients, cross-sectionnally and longitudinally for 14 of them.

## Patients and Methods

We analyzed the IS and serum of 35 patients with SSc in comparison to 30 HV. The diagnosis of SSc was made according to the international recommendations of the 2013 ACR/Eular for the classification of systemic sclerosis [31] and the distinction of the cutaneous forms into limited and diffuse according to the classification of Leroy et al. in 1988 [32]. Early forms of SSc, also called limited, have been classified according to Leroy and Medsger [33] while sine scleroderma fulfilled Poormoghim criteria [34]. The protocol was approved by ethics committee of CHU Liège, and all subjects gave written consent before their enrolment (Belgian number: B707201422832; ref 2014/302).

### Sputum induction and processing

The technical process used is the same as previously described in our methodological manuscript [24]. After premedication with 400 □g inhaled salbutamol, sputum was induced by inhalation of hypertonic (NaCl 5%) or isotonic (NaCl 0.9%) saline according to the FEV-1 value (> or ≤ than 65% predicted). Saline was combined with additional salbutamol delivered by an ultrasonic nebulizer (Ultra-Neb 2000; Devilbiss, Somerset, PA, USA) with an output set at 0.9 ml/min. Each subject inhaled the aerosol for three consecutive periods of 5 min for a total of 15 min. For safety reasons, FEV-1 was monitored throughout the induction and it was stopped if FEV-1 fell by more than 20% from baseline. The whole sputum was collected in a plastic container, weighed, and homogenized by adding 3 volumes of phosphate-buffered saline (PBS), vortexed for 30 s, and centrifuged at 800 g for 10 min at 4 □C. Supernatant was separated from cell pellet and stored at − 80 □C. The cells were resuspended in a solution containing 5mM dithiothreitol without Ca^++^ and Mg^++^ and gently rocked for 20 min at room temperature. The cell suspension was then centrifuged again at 400 *g* at 4 □C for 10 min. Squamous cells, total cell counts and cell viability checked by trypan blue exclusion were performed with a manual hemocytometer [27]. The differential leukocyte count was obtained using a cytospin stained with Rapi Diff II (Atom Scientific, Manchester UK) on 500 non squamous cells. Ten SSc patients and 5 HV were unable to realized their induced sputum (induction failure). Therefore only 25 SSc patients and 25 HV were considered for analysis in our study.

### Biomarkers measurements in the IS and serum

We chose several molecules known to be of interest in inflammation, remodeling and fibrotic processes of IPF (growth factors, cytokines, matrix metalloproteases -MMPs), and determined their levels in the IS and in the serum [35]. The concentration of TNF-α, MMP-7, IL-8, IGFBP-1 and YKL-40 were assessed by multiplex using Fluorokine1 Multianalyte Profiling Kits (R and D Systems, Minneapolis, MN, USA) according to the manufacturer’s instructions. The detection limit for the assays were 4-230-3-170 and 200 pg/ml, respectively. The concentration of the other proteins were measured by enzyme-linked immunosorbent assay (ELISA) using DuoSet kit (R and D Systems): TGF-β, MMP-9, IGFBP-2 and IGFBP-3. The detection limits for these kits were 7-25-30-130-450 pg/ml, respectively.

### Pulmonary function tests

Lung function tests were performed in our routine respiratory laboratory at CHU Liège. All spirometric tests performed were measured using the pneumotachograph JaegerMasterlab system (Erich Jaeger GmbH, Wuzburg, Germany). The expiratory volume in one second (FEV-1) and forced vital capacity (FVC) were measured in accordance with the recommendations of the European Respiratory Society (ERS)[36]. Results were expressed in ml and as percentage of predicted normal values. The Tiffeneau index or FEV-1/FVC is expressed as percentage. The diffusion capacity of CO (DLCO) and carbon monoxide transfer coefficient (KCO or DLCO/VA) were measured by the single breath testing technique (Sensor Medics 2400 He / CO Analyzer System, Bilthoven, The Netherlands).

### Statistical analysis

When the data showed normal distribution, they were compared with a one-way ANOVA, followed by Bonferoni post-hoc test. When the data did not showed a normal distribution, they were compared by Kruskall-Wallis followed by Dunns post-hoc test. Correlations between variables were assessed using Spearman’s rank correlation test. A p<0.05 was considered as significant. The statistical analysis and graph were performed with Graph Pad Prism 5 software. To evaluate correlations between biomarkers and PFT, we realized simple non-parametric Spearman method. In order to normalize the longitudinal analysis, the longitudinal evolution was calculated by performing the subtraction of the first PFT (reference value) and the value at the time of the second visit (day of sputum induction) divided by the number of years between the two visits.

## Results

### Subject demographic and functional characteristics including lung function

The demographic, functional and treatment characteristics of the subjects are given in Table 1.

**Table 1.**
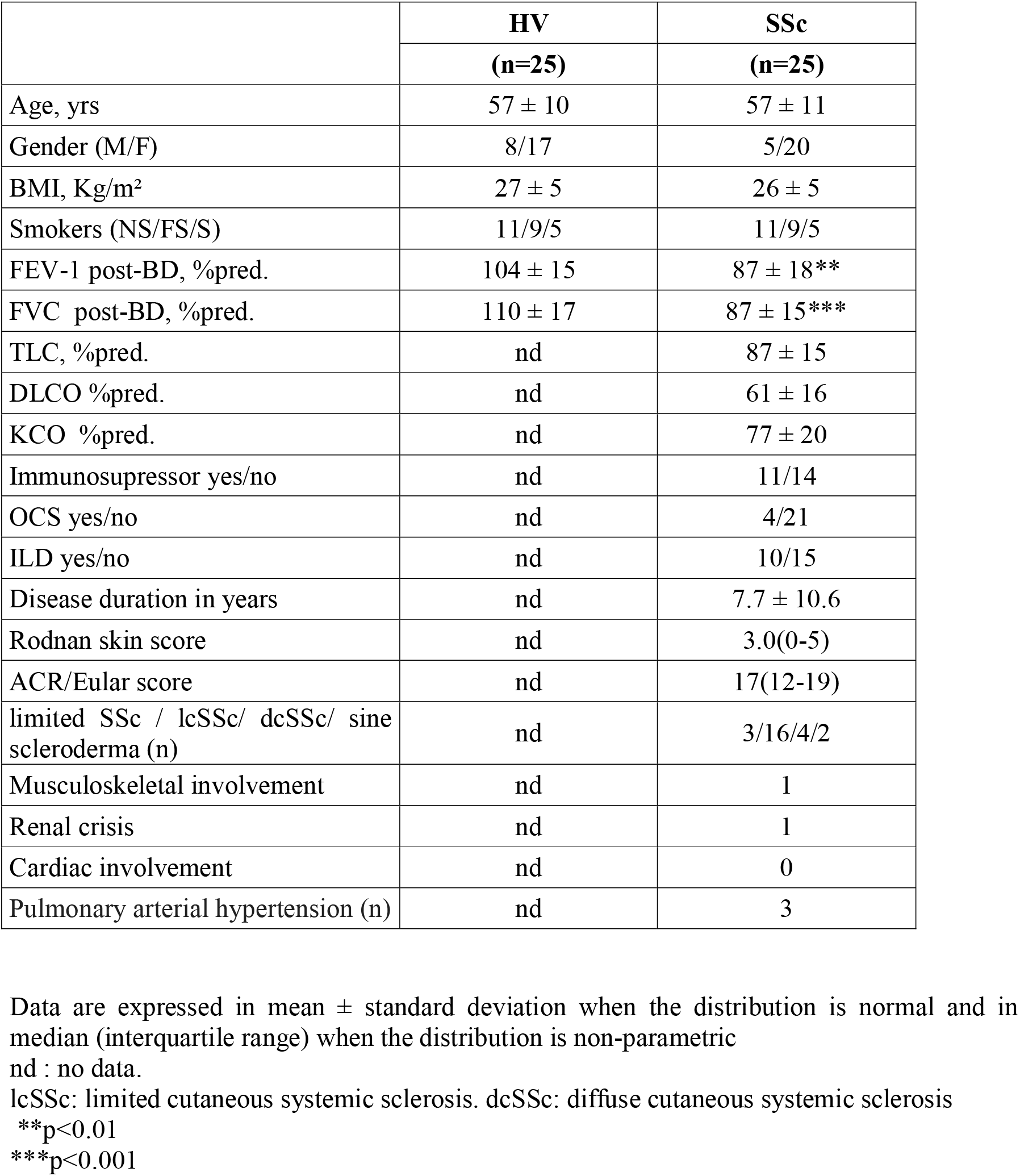
Subjects characteristics.

50 participants were finally included in this study: 25 SSc patients and 25 HV. The SSc population was composed of 3 limited SSc (12%), 16 lcSSc (64%), 4 dcSSc (16%) and two sine scleroderma (8%). Among SSc patients, 6 lcSSc and 4 dcSSc had ILD. HV group was similar to SSc group except for lung function with FEV-1 (p<0.01) and FVC (p<0.001) significantly lower in SSc patients compared to HV. SSc patients also exhibited reduced DLCO and KCO with a preserved TLC (Table 1). Mean (SD) disease duration, calculated from the onset of the first non-Raynaud symptom was 7.7 ± 10.6 years. Mean (SD) clinical scores were 3.0 (0-5) for the Rodnan score and 17 (interquartile 12-19) for the ACR/Eular score (Table 1). Five subjects had a restrictive pattern on PFT with a mean TLC of 67%. Three of the SSc patients had PAH. At the time of evaluation, 11 patients (44%) were receiving immunosuppressive therapy (1 association of azathioprine (AZA)/oral corticosteroid (CS), 1 hydroxychloroquine (HCQ)/ mycophenolate mofetil (MMF)/CS, 2 MMF/CS, 1 HCQ, 1 methotrexate (MTX), 1 AZA, 1 MMF, 1 HCQ/MTX, 1 advagraf/AZA/CS and 1 MMF/rituximab/CS.

### Sputum cellularity and supernatant biomarkers

The IS weight of SSc patients was significantly lower than in HV (p<0.01), the cellularity being however similar in both groups (Table 2) with a predominance of macrophages and neutrophils. In the SSc group, IGFBP-1 (p<0.0001), TGF-β (p<0.05), IL-8 (p<0.05), YKL-40 (p<0.0001) and MMP-7 (p<0.01) levels in the supernatant of IS were increased compared to HV whereas no difference was found in the levels of IGFBP-2 and of MMP-9 (Figure 1). IGFBP-3 and TNF-□ were not detectable. No difference was observed in these parameters between SSc-ILD and SSc-nonILD patients (data not shown).

**Table 2.**
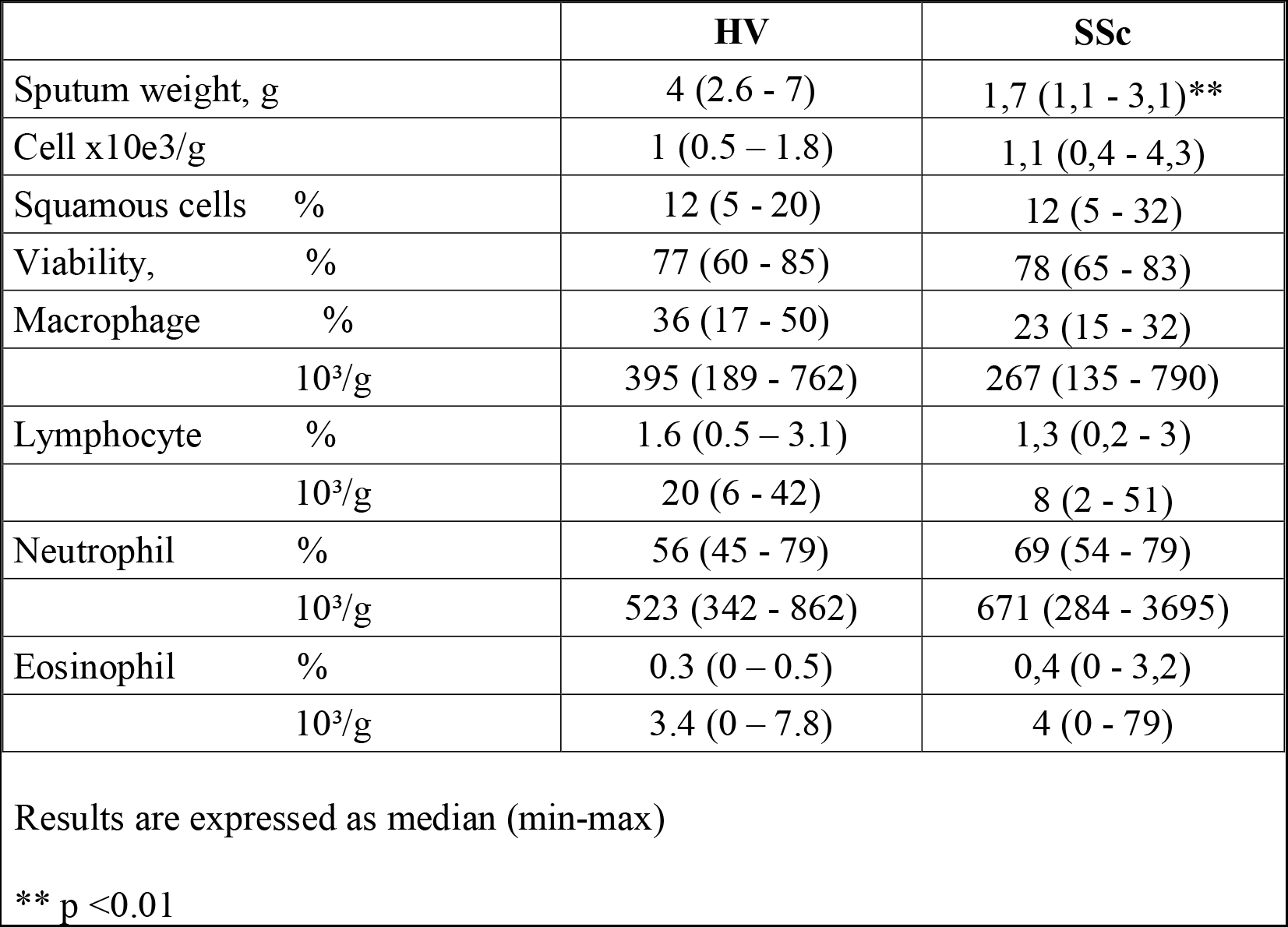
Sputum cell counts.

|  | **HV** | **SSc** |
| --- | --- | --- |
| Sputum weight, g | 4 (2.6 - 7) | 1,7 (1,1 - 3,1)** |
| Cell x10e3/g | 1 (0.5 – 1.8) | 1,1 (0,4 - 4,3) |
| Squamous cells % | 12 (5 - 20) | 12 (5 - 32) |
| Viability, % | 77 (60 - 85) | 78 (65 - 83) |
| Macrophage % | 36 (17 - 50) | 23 (15 - 32) |
| 10<sup>3</sup>/g | 395 (189 - 762) | 267 (135 - 790) |
| Lymphocyte % | 1.6 (0.5 – 3.1) | 1,3 (0,2 - 3) |
| 10<sup>3</sup>/g | 20 (6 - 42) | 8 (2 - 51) |
| Neutrophil % | 56 (45 - 79) | 69 (54 - 79) |
| 10<sup>3</sup>/g | 523 (342 - 862) | 671 (284 - 3695) |
| Eosinophil % | 0.3 (0 – 0.5) | 0,4 (0 - 3,2) |
| 10<sup>3</sup>/g | 3.4 (0 – 7.8) | 4 (0 - 79) |

Results are expressed as median (min-max)
\*\* p <0.01

**Figure 1.**
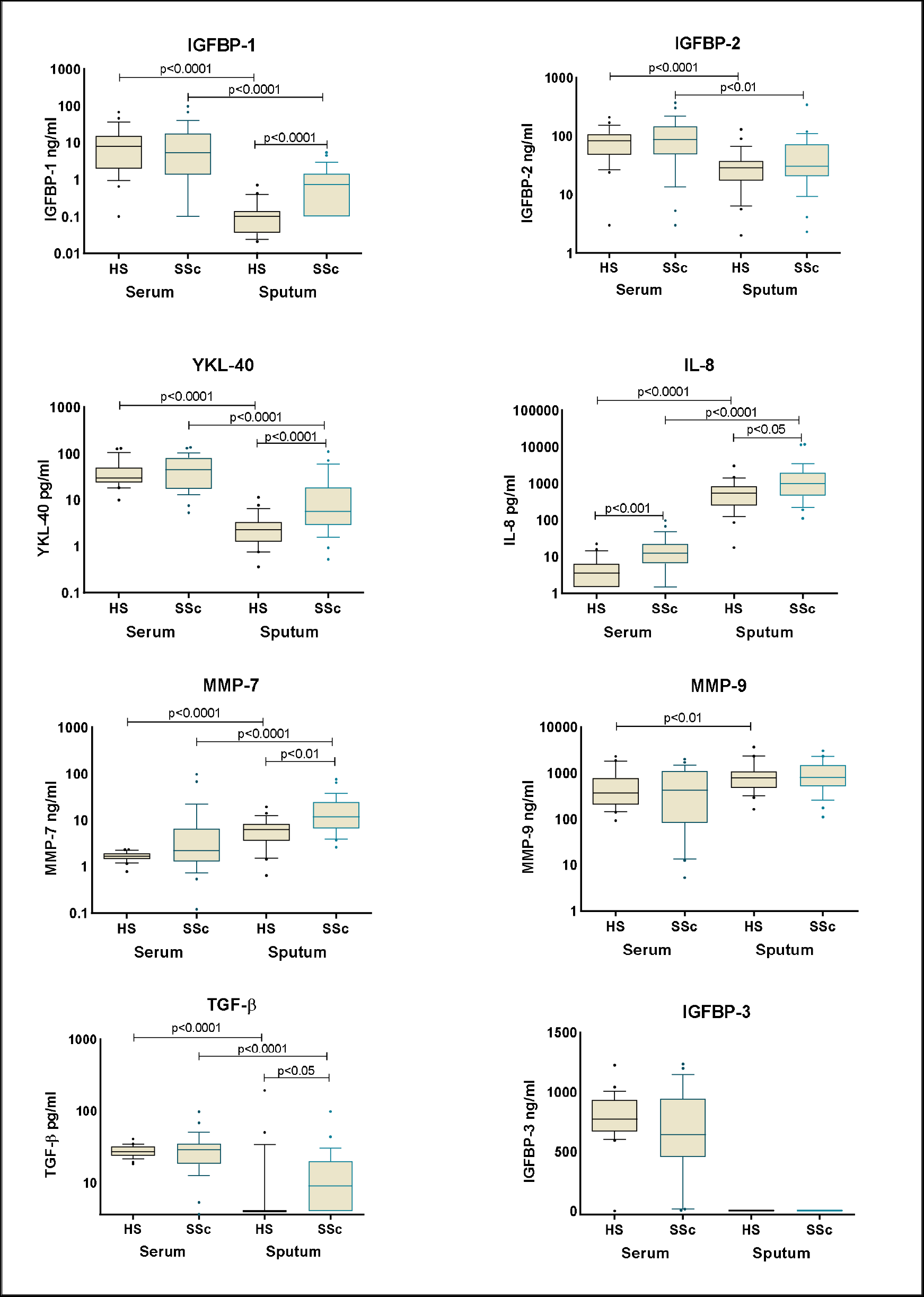
Comparison serum/sputum biomarkers.

### Correlations between sputum biomarkers and lung function

Correlations were observed between IS IL-8 levels and FEV-1 (%)(r=-0.53, p<0.01) and FVC (%) (r=-0.51,p<0.01) (Table 3 and Figure 2). Further, in 14 SSc patients, 2 longitudinal PFT were obtained allowing the determination of the annualized variation (□) of the tested parameters (see statistical analysis). Correlations were then also observed between IS IL-8 levels and □KCO (%) (r=0.57, p<0.05), between IS TGF-□ levels and □FEV-1 (%) (r=-0.57, p<0.05), and between IS IGFBP-2 levels and □KCO (%) (r=0.56, p<0.05) (Table 3 and Figure 2). All observed correlations were lost when applied only to SSc-ILD patients (data not shown).

**Table 3.**
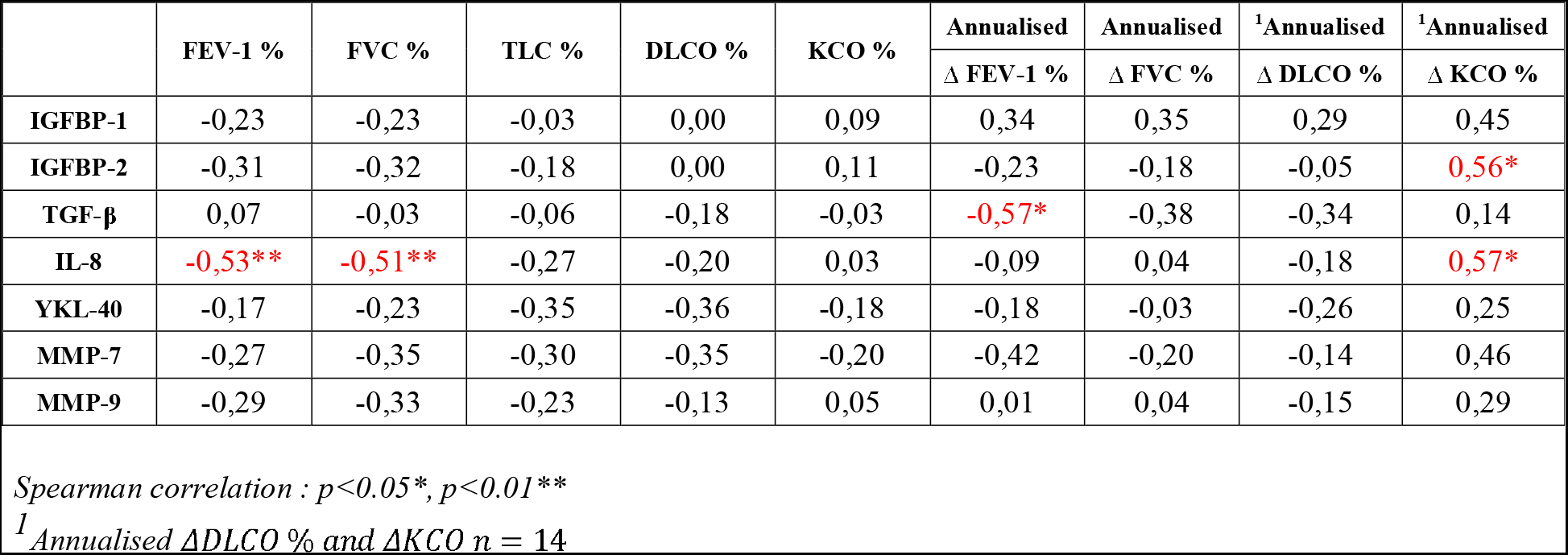
Correlation between sputum biomarkers and lung function in SSc group.

**Figure 2.**
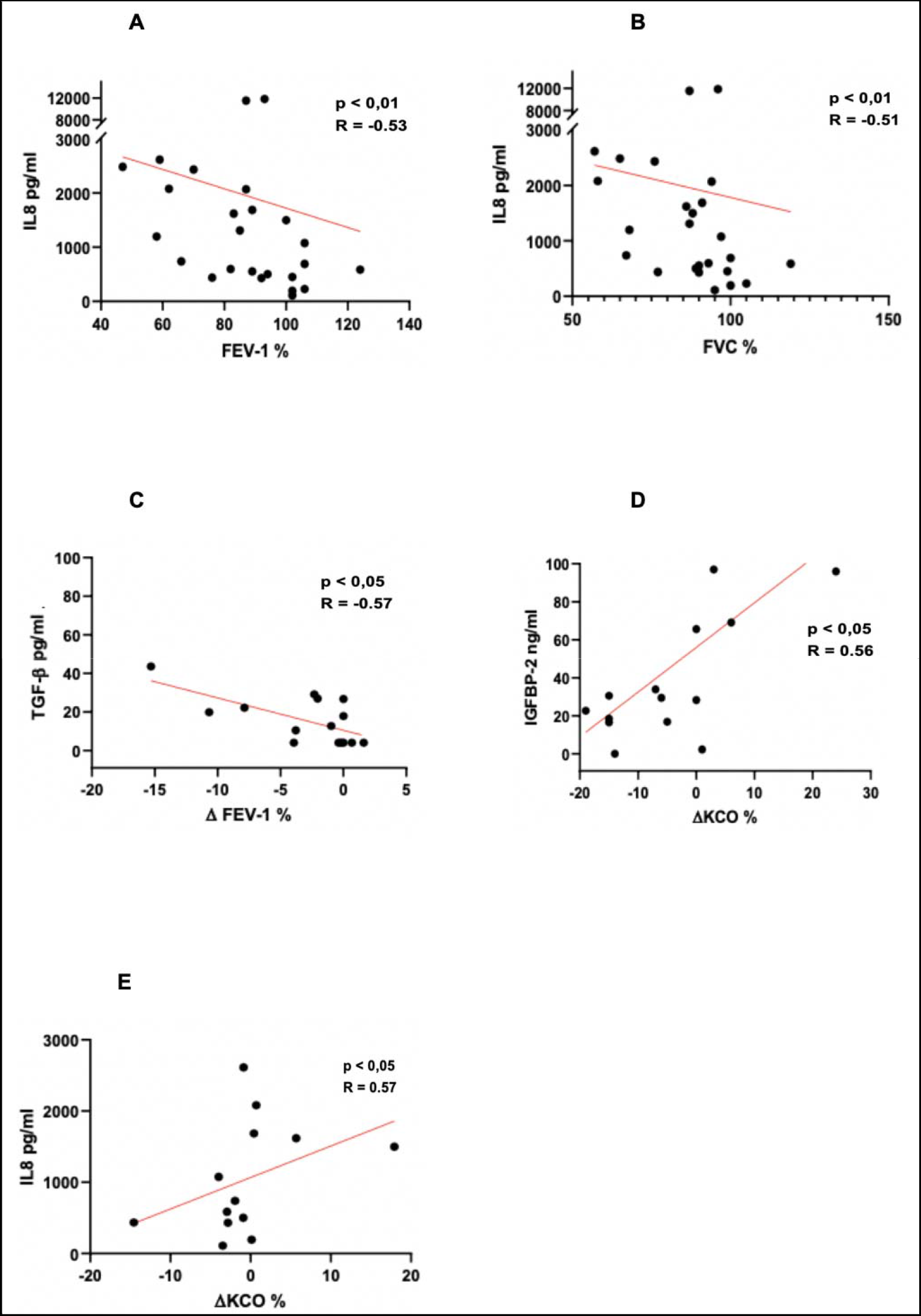
Correlation between sputum biomarkers and PFT.

### Correlations between sputum biomarkers and clinical scores

Correlations were also found between IS IGFBP-2 and the Rodnan score (r=0.46, p<0.05) and between IS IGFBP-2 (r=0.43), YKL-40 (r=0.41) or MMP-7 (r=0.44) and the ACR/EULAR classification criteria (all p<0.05). No correlation was observed between BMI or duration of disease at baseline with all parameters tested. Data are shown in table 4.

**Table 4.**
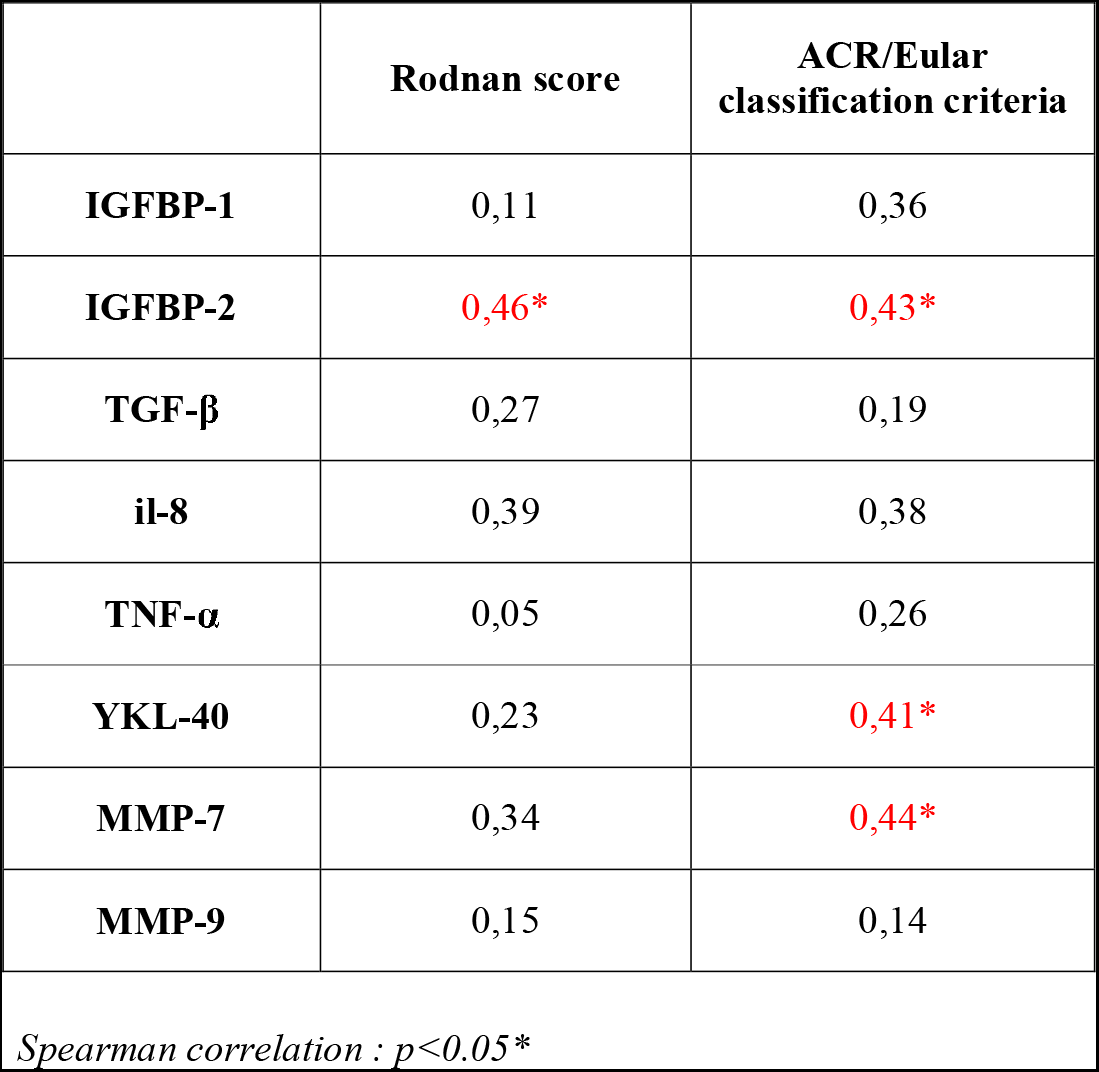
Correlation between sputum biomarkers and clinical scores in SSc group.

|  | Rodnan score | ACR/Eular classification criteria |
| --- | --- | --- |
| **IGFBP-1** | 0,11 | 0,36 |
| **IGFBP-2** | **0,46\*** | **0,43\*** |
| **TGF-β** | 0,27 | 0,19 |
| **il-8** | 0,39 | 0,38 |
| **TNF-α** | 0,05 | 0,26 |
| **YKL-40** | 0,23 | **0,41\*** |
| **MMP-7** | 0,34 | **0,44\*** |
| **MMP-9** | 0,15 | 0,14 |

Spearman correlation : $p < 0.05^*$

### Serum biomarkers

Among all the parameters evaluated, only serum IL-8 levels were significantly higher in SSc patients compared to HV (p<0.001) (Figure 1). There was also no difference between the SSc-ILD and SSc-nonILD patients (data not shown). TNF-□ remained undetectable in both groups.

### Comparison between sputum and serum

If we now compare tested parameters in IS and in serum, we have observed that IGFBP-1, IGFBP-2, TGF-□ and YKL-40 levels were lower in the IS of HV compared to serum values, that IL-8, MMP-7 and MMP-9 levels were higher. Applied to the SSc population, we found exactly the same profile, except MMP-9 levels not different between IS and serum (Figure 1). The only correlation found was between IS and serum was for MMP-7 (r=0.50, p<0.01) and YKL-40 (r=0.57, p<0.01) in HV.

## Discussion

In this study, we report for the first time, in the IS of patients with SSc compared to HV, a significant increase of several proteins, IGFBP-1, TGF-β, IL-8, YKL-40 and MMP-7 known to be also elevated in the IS of IPF patients. No difference was found in the levels of IGFBP-2 and MMP-9, whereas IGFBP-3 and TNF-□ were undetectable in both groups.

Pulmonary involvement is a determining factor of morbidity and mortality in systemic sclerosis. For this purpose, the importance of early diagnosis and rapid management of complications is primordial while limiting invasive techniques. The use of IS is well established in detecting alveolitis secondary to several inflammatory lung diseases but its role in the assessment of SSc patients is not yet well defined [23].

In our study, we have found a rate of induction failure of 28.5% in SSc patients and of 17% in HV, rather similar to the classical rate of 15% in asthmatic patients and 20% in HV [24]. Whereas the weight of the IS of SSc patients was lower than that of the IS of HV, we did not observe any significant difference in terms of cellularity and viability. Contrasting results have been previously reported. Damjanov et al. [26] studying the IS of 68 SSc-ILD patients (versus 18 HV) observed a significant increase in total number of inflammatory cells, of neutrophils, lymphocytes and eosinophils, and lower macrophages, in SSc patients. Yialmaz et al. [27] studying the IS of 17 SSc-ILD patients (versus 12 non-ILD pulmonary controls) observed a significant increase in total number of inflammatory cells, of lymphocytes and macrophages, and lower neutrophils, in SSc patients. Litinsky et al. [25] studying the IS of 20 SSc-patients (versus 16 HV) reported an isolated decrease in lymphocytes (p=0.045), with no change in the percentage of neutrophils, macrophages and eosinophils. This heterogeneity of results might be related to the presence of concomitant immunosuppresive therapies, 44% of our patients, 45% in Litinsky’s work and only in 25% of Damjanov’s one, but also to the type of controls chosen. Our observation is therefore close to the findings of Litinsky et al. [25].

The transforming growth factor beta (TGF-β) is well known for its role in fibrosing process. TGF-β is a stimulus for pulmonary fibrogenesis by its activity in the control of the remodeling of the extracellular matrix. TGF-β is also known to be involved in the fibroblast activity [37]. As expected, TGF-β was found increased in IS of SSc patients without any difference between SSc-ILD and SSc-nonILD patients. In that context, we can hypothesize that the fibroblastic responsiveness under TGF-β stimulation could vary between patients suffering from SSc inducing a personal susceptibility to develop ILD partly induced by genetic, environmental and inflammatory factors [38-40]. Interestingly, we found a negative correlation between IS TGF-□ levels and ΔFEV-1%. This is probably why, specific anti-fibrotic therapies focusing on TGF-β pathway (pirfenidone) could be possibly used in SSc-ILD. The LOTUSS study published in 2014 evaluated the potential of pirfenidone in SSc (safety study). Nevertheless, this one was not designed to evaluate its efficacy [41].

The matrix metalloproteinase MMP-7 levels were also higher in the IS of SSc patients compared to HV. MMP-7 was previously reported by Huh et al. [42] to be increased in the BALF of IPF patients, and we recently described a raise of MMP-7 both at gene and protein levels in the IS from IPF compared to COPD patients and HV [23], correlated to PFT [43]. MMP-7 levels are known to be increased both in IPF and SSc-ILD patients compared to HS [44]. Due to the heterogeneity of our cohort with regard to the limited number of patients with ILD, we didn’t found any difference between those and HV. Moreover, its serum level is more increased in SSc-ILD than in SSc-nonILD and in patients with higher severity (dyspnea and DLCO levels below 60%) [45]. As this observation is also found in IPF, we can hypothesize that this protein takes part of the lung fibrosing process found both in IPF and SSc-ILD. In the same line, a recent study (Senscis) identified that nintedanib known to be effective in IPF reduces lung function decline in SSc-ILD patients [46,47].

The IGFBP-1 protein is part of a group (IGFBPs) known to be involved in the regulation of insulin-like growth factor (IGF). IGF-1 (and IGF-2) play an important role in the growth, differentiation and cellular metabolism [49, 50]. We have previously shown that IGFBP-1 and IGFBP-2 proteins were raised in serum [51] while IGFBP-2 was increased in sputum of IPF [23]. The increased levels of sputum IGFBP-1 reinforces the potential role of IGFBPs in SSc. Supporting the role of IGFBPs in disease progression, we interestingly found a positive correlation between IGFBP-2 levels and KCO annualized variation which is a specific marker of alveolo-capillary dysfunction. In this study we observed a clear increase of IGFBP-1 IS levels whereas its level is known to be very low in HV as well as in COPD and IPF patients [23].

YKL-40, a member of the mammalian chitinase-like protein family, is produced by various cell types, including macrophages, neutrophils, monocytes, airway epithelium cells, vascular smooth muscle cells, synovial cells, chondrocytes, and breast cells [52]. Elevated blood YKL-40 levels have been observed in various diseases characterized by inflammation, abnormal cell growth, and tissue remodeling, such as cardiovascular disease, diabetes, cancer, and idiopathic pulmonary fibrosis [53-55]. Tong et al. suggested that YKL-40 might be implicated in bronchial inflammation and remodeling in COPD and be considered as a useful biomarker for COPD diagnosis and monitoring [56]. In our study, SSc patients exhibited higher IS levels than HV. This is an original finding as the protein had not been measured in the airway/lung compartment of patients suffering from SSc. Conversely to results BALF levels of YKL-40 are known to be increased in multiple fibrosing lung diseases [57-59], our previous studies failed to identify any significant increase in sputum of IPF patients [23]. Interestingly, IS and serum levels of YKL-40 and of MMP-7 are positively correlated in HV, correlations surprisingly lost in SSc patients suggesting intense local production in the bronchioloalveolar tract.

IL-8, a well-known chemotactic agent for neutrophils [59], has already been shown as increased both at gene and at protein level in the IS of IPF [23, 60, 61] and of COPD patients [23] compared to HV. Higher BALF IL-8 levels have been previously reported in SSc-ILD patients as compared to SSc-nonILD patients, to idiopathic ILD patients or to nonILD HV [62]. Further, they correlated negatively with functional lung parameters TLC, FVC and DLCO [62]. We observed elevated IS IL-8 levels compared to HV, as well as negative correlations with FEV-1%, FVC% and positive correlation with □KCO%, therefore in agreement with others suggesting IL-8 to be a prognostic marker related to the severity of the disease [62]. Serum studies also indicated that IL-8 only was higher in SSc patients compared to HV, but at concentration almost 100 times less then IS levels suggesting a high biomarker production in the bronchioloalveolar tract.

Correlations between IS profile and lung functional parameters strongly support a role for these parameters in the pathophysiology of the disease, and particulary of bronchoalveolar dysfunction even if clinical ILD is not documented. It will be recalled that no difference was evidenced in any parameter between SSc-ILD and SSc-nonILD patients. Further, not all SSc patients had pathological PFT: 7 (5 ILD) with FEV-1% < 80%, 6 (5 ILD) with FVC1%<80%) and 15 (9 ILD) with DLCO<70% of predicted values. It is therefore tempting to suggest that IS profiles in SSc patients, by similarities to those in IPF patients, might represent an infraclinical inflammatory and fibrotic lung process. Cross-sectional negative correlations with FEV-1% (IL-8) and with FVC% (IL-8) and longitudinal negative correlations with FEV-1% (TGF-□) support this hypothesis. It is lastly of strong interest to also observe significant positive correlations between IS IGFBP-2, MMP-7 or YKL-40 levels and the ACR/Eular classification criteria linking IS levels of these parameters to the global extension of the disease. Similarly, the significant positive correlation between IS IGFBP-2 levels and the modified Rodnan skin score suggest a role of this parameters in the thickness and the diffusion of the skin sclerosis process.

Finally, when compared to the same parameters identified in the IS of IPF patients [23], it is interesting to note that there exist similarities between both diseases reflected by elevated TGF-□, IL-8 and MMP-7 levels, but also difference: IGFBP-1 and YKL-40 levels are elevated in SSc patients and not different from HV in IPF; IGFBP-2 and MMP-9 levels are similar to HV in SSc patients and higher in IPF patients and IGFBP-3 and TNF-□ are present in IPF patients but not in SSc ones. This indicates that both diseases do not share the same whole pathophysiological process. As aforementioned, one of the potential limitation of the study is the cross sectional design that precludes any interpretation of the predictive values of these biomarkers in the follow-up of the patients. Nevertheless, the clinical follow-up as well as the PFT reevaluation led us to rise some hypothesis of the predictive value of each biomarker.

## Conclusion

Our data showed a clear increase in the levels of several critical growth factors, matrix metalloproteinases and chemokines in IS of SSc patients. The sputum analysis could become a suitable and less invasive tool than BALF to predict and monitor evolution of the disease and treatment response should be investigated with a larger prospective longitudinal trial. Correlations between IS biomarkers and lung functional parameters or parameters that reflect the global diffusion or skin severity in SSc patients give clinical relevance to the sputum profile observed in this disease. IS composition of SSc patients mimick that of IPF patients, but is not identical with probably own original pathways.

## Data Availability

I declare that all data in this manuscript is available

## Acknowledgments

Thank you to the FIRS (Fonds d’Investissement de la Recherche Scientifique) from CHU Liège for their funding in our study and all co-authors for their contribution:

**Conceptualization:** PJ, JG, MH, RL, MM

**Data curation:** PJ

**Formal analysis:** PJ, MH, JG

**Investigation:** JG, MH

**Methodology:** MH

**Project administration:** JG

**Supervision:** MM, RL, JG

**Validation:** RL, MM

**Visualization:** PJ, BA, CM, DS

**Writing – original draft:** PJ

**Writing – review & editing:** PJ, JG

## Supporting information

